# Gender Differences in Treatment Burden and Care Sustainability Among Adults Living With Diabetes

**DOI:** 10.64898/2026.09.08.26362531

**Authors:** Misk Al Zahidy, Francisco D. Rivadeneira, Omar M. Rabeaa, Brandon N. Shah, Megan E. Branda, Juan P. Brito, Victor M. Montori

## Abstract

**Objective:** Women carry disproportionate shares of caregiving and other life demands that may compound the work of diabetes self-management. We sought to examine gender differences in overall and digital treatment burden and their key contributors.

**Research Design and Methods:** Cross-sectional analysis of two cohorts of adults with type 1 or type 2 diabetes attending outpatient endocrinology visits who completed the Treatment Burden Questionnaire Plus Digital (TBQ+D) (n=472). Multivariable regression estimated gender differences in overall and digital treatment burden, ability to live a normal life, and sustainability of current self-care effort, adjusting for age, diabetes type, insulin intensity, HbA1c, and cohort. Item-level analyses identified contributors to observed differences.

**Results:** Compared to men, women reported higher median overall treatment burden (33 vs 20, adjusted mean difference 10.6 points, 95% CI 5.1–16.3; *p*<.0001) and digital treatment burden (13 vs 7, adjusted mean difference 3.5 points, 95% CI 1.1–6.1; *p*=.009). They also had higher odds of not living a normal life (OR 1.77, 95% CI 1.18–2.67; *p*=0.006) and difficulty sustaining current self-care efforts indefinitely (OR 2.01, 95% CI 1.10–3.66; *p*=0.023). Digital burden differences were concentrated in annoyance, precautions, and time and effort required to use digital medicine tools, whereas contributors to differences in overall burden varied, with relationships, self-monitoring, and injections contributing most.

**Conclusions:** After adjusting for treatment intensity and other factors, women living with diabetes were twice as likely to be overwhelmed by diabetes care than men. Their greater vulnerability to treatment burden demands healthcare service redesign and individualization of plans of care that minimally disrupt patients’ and caregivers’ lives.

**ARTICLE HIGHLIGHTS:**

- **Why did we undertake this study?**

Women may experience greater treatment burden across chronic conditions, reflecting competing (caregiving) demands that reduce capacity for self-management. Gender differences in diabetes, especially digital burden, remain unclear.

- **What is the specific question we wanted to answer?**

Do women with diabetes experience greater overall and digital treatment burden than men?

- **What did we find?**

Women reported greater overall and digital burden and more difficulty living a normal life and sustaining care than men. Differences were larger for digital annoyance, precautions, and effort, plus relationships, self-monitoring, and injections.

- **What are the implications?**

Treatment burden cannot be inferred from treatment intensity alone. Diabetes care should consider whether treatment work fits within each patient’s life and can be sustained. Women with diabetes may be particularly vulnerable to becoming overwhelmed by the work of diabetes care.

---

Diabetes management requires substantial work from patients between visits, including taking medications, monitoring glucose, adjusting insulin or other treatments, planning meals and physical activity, attending appointments, completing laboratory testing, and coordinating care.(1) As digital medicine tools have become increasingly integrated into diabetes care, patients may also need to respond to alerts and alarms, troubleshoot devices, manage supplies and software, and integrate technologies such as continuous glucose monitors, insulin pumps, and companion applications into daily life.(2) The effort required to manage this care, and the effect that effort has on patients’ lives, is commonly described as treatment burden. Treatment burden reflects the balance between the workload imposed by care and a patient’s capacity to manage it, as described in the Cumulative Complexity Model(3), and is central to minimally disruptive medicine, which calls for care that fits within patients’ lives rather than adding unnecessary work.(4) When the work required by care exceeds what patients can reasonably manage, treatment burden may interfere with daily life, make care difficult to sustain, and contribute to poorer adherence and health outcomes.(5; 6)

Because treatment burden reflects the relationship between the work required by care and a person’s capacity to manage that work, it may not be experienced equally across patients. Women have repeatedly been reported to experience greater treatment burden across chronic conditions.(6–9) Qualitative studies suggest several possible contributors, including caregiving responsibilities, competing family and work demands, and structural or financial barriers to care, all of which may reduce the time, energy, and resources available to manage treatment.(10–16) In diabetes, these demands may be particularly important because self-management often requires continuous attention to medications, glucose monitoring, devices, and treatment decisions. Recent work has also highlighted sex- and gender-related differences in diabetes management as an understudied area, particularly in relation to self-care and mental health burden.(17) What remains unclear is how these gender differences are reflected in the day-to-day work of diabetes care. Existing studies have also generally focused on total treatment burden scores, providing limited insight into which specific aspects of care, including digital tool use, contribute most to these differences.

Using the Treatment Burden Questionnaire Plus Digital (TBQ+D)(2; 18; 19), we examined gender differences in overall and digital treatment burden among adults living with diabetes and identified the aspects of treatment that contributed most to the observed differences. To understand further the impact of the burden of treatment, we also examined how participants assessed the sustainability of their self-management efforts and how this burden disrupts their ability to live a normal life, exploring whether these differ by gender.

## RESEARCH DESIGN AND METHODS

### Design, Setting, and Participants

We conducted a cross-sectional analysis pooling two cohorts of adults aged 18 years or older living with type 1 or type 2 diabetes who were recruited within the Mayo Clinic Division of Endocrinology, Diabetes, Metabolism, and Nutrition in Rochester, Minnesota, under separate institutional review board–approved protocols. The first cohort was recruited from May 2024 to October 2024 as part of the validation of the Treatment Burden Questionnaire Plus Digital (TBQ+D) (IRB #23-007631)(19), and the second, the Encounters cohort, was recruited from January 2025 to May 2026 as part of a longitudinal study of patient-clinician encounters (IRB #24-012956).(20) Both cohorts included adults attending an in-person outpatient endocrinology visit who completed a post-visit survey that included the TBQ+D. The questionnaire was self-administered electronically, with a paper version available if preferred. Gender was obtained from the study data source used for each cohort and was self-reported in the electronic health record.

This study is reported in accordance with the Strengthening the Reporting of Observational Studies in Epidemiology (STROBE) guidelines.(21)

### Measure

Treatment burden was assessed using the Treatment Burden Questionnaire Plus Digital (TBQ+D), a validated patient-reported measure that extends the original Treatment Burden Questionnaire to capture the work associated with digital medicine tools.(2; 18; 19) The TBQ+D includes 18 core treatment-burden items and 6 digital treatment-burden items, each scored from 0 (not a problem) to 10 (big problem), yielding an overall treatment burden score ranging from 0 to 180 and a digital treatment burden score ranging from 0 to 60.

Only participants who reported using digital medicine tools completed the digital burden items; therefore, analyses of digital treatment burden were restricted to participants using these tools.

Two additional items assessed whether participants felt able to live a normal life and whether they believed they could continue investing the same amount of time, energy, attention, and money in their health indefinitely. These items were analyzed as separate binary outcomes.

### Data Analysis

We used descriptive statistics to summarize participant characteristics by gender. Continuous variables are presented as means and standard deviations and categorical variables as frequencies and proportions. Differences between women and men were assessed using two-sided t tests or chi-square tests, as appropriate.

We examined gender differences in overall and digital treatment burden using generalized linear models with a Gaussian distribution and log link and differences in the ability to live a normal life and sustain the current level of self-care effort using multivariable logistic regression. Models were adjusted for age, diabetes type, insulin intensity, HbA1c, and study cohort (Validation vs. Encounters).

We explored whether the association between age and overall treatment burden differed by gender and whether the association between gender and overall treatment burden differed by diabetes type using age-by-gender and gender-by-diabetes type interaction terms, respectively, in adjusted regression models evaluated using Wald tests.(22) We also estimated the association between age and overall treatment burden separately for women and men using the same adjustment variables. We also examined which individual TBQ+D items contributed most to the observed gender differences. Because domain scores are the sum of their individual items, we decomposed the unadjusted mean difference between women and men into the contribution of each item. We then re-estimated the gender difference for each item using generalized linear models with a Gaussian distribution and log link, adjusted for age, diabetes type, insulin intensity, HbA1c, and study cohort (Validation vs. Encounters). These covariates were selected to account for clinically relevant differences in diabetes and treatment characteristics that may be associated with treatment burden, while cohort was included to account for differences between the two independently recruited study samples.

We also examined overall treatment burden using two anchor-based thresholds. First, we applied the previously published Patient Acceptable Symptom State (PASS) threshold for treatment burden, derived using an anchor assessing whether patients could sustain their current investment of time, energy, and money in health care over the long term.(23) The PASS corresponded to 39% of the maximum possible TBQ score (59 of 150 points), with scores at or above this threshold indicating potentially unacceptable or unsustainable treatment burden. First, we characterized the cohorts using the absolute PASS cutoff (i.e., 59 points) only on the overall TBQ score and the relative cutoff (i.e., 39%) on the overall TBQ+D (i.e., 70 points on a maximum range of 0 to 180). Because this latter PASS threshold has not been validated for the TBQ+D, the rescaled 70-point threshold was considered exploratory. Second, we estimated a sample-specific threshold using the study’s sustainability item as the anchor, reproducing the approach used to generate the original absolute PASS TBQ cutoff.(23)

Primary analyses used complete cases for the variables required for each model; no missing-data imputation was performed. As a sensitivity analysis, we repeated the adjusted analyses separately within each cohort (Validation vs. Encounters). All analyses were conducted using Python (version 3.12.3), with statistical significance defined as a two-sided *p*<.05.

## RESULTS

### Baseline Characteristics

Among the 472 participants in the final analytic sample, 469 had gender recorded as woman or man and were included in gender-comparison analyses; two participants identified as another gender, and one had missing gender data. Of the 469 participants included in gender-comparison analyses, 229 were women and 240 were men.

Women and men were similar in age, HbA1c, diabetes type, cohort composition, and use of digital medicine tools **(Table 1)**. Intensive insulin use was more common among women than men (n=167 (73%) vs n=142 (59%); *p*=.002). Overall treatment burden was higher in participants with type 1 than type 2 diabetes in unadjusted analyses (median 32 vs 23; *p*=.0016).

**Table 1.** Baseline characteristics of participants.

| Characteristic | Women<br>(n=229) | Men<br>(n=240) | *P* value |
| --- | --- | --- | --- |
| Age, mean (SD), years* | 55.8 (16.5) | 57.4 (16.5) | 0.29 |
| HbA1c, mean (SD), %* | 7.4 (1.5) | 7.7 (2.0) | 0.09 |
| (mmol/mol) | [57.7 (16.4)] | [60.8 (21.6)] |  |
| Missing, n (%) | 2 (0.9) | 5 (2.1) |  |
| Type 1 diabetes, n (%)† | 118 (51.5) | 102 (42.5) | 0.06 |
| Intensive insulin use, n (%)† | 167 (72.9) | 142 (59.2) | 0.002 |
| Validation cohort, n (%)† | 151 (65.9) | 147 (61.3) | 0.34 |
| Digital medicine tool use, n (%)† | 207 (90.4) | 206 (86.2) | 0.21 |
| Missing, n (%) | 0 (0.0) | 1 (0.4) |  |

\*Compared using a two-sided t test.
†Compared using a chi-square test.

### Adjusted Gender Differences in Treatment Burden

Women reported higher unadjusted median overall treatment burden than men (33 vs 20) and higher digital treatment burden (13 vs 7) **(Figure 1).** After adjustment for age, diabetes type, insulin intensity, HbA1c, and cohort (Validation vs. Encounters), women had an overall treatment burden score that was 10.6 points higher than men (95% CI 5.06–16.31; *p*<.0001; n=436) and a digital treatment burden score that was 3.5 points higher (95% CI 1.13–6.09; *p*=.009; n=402) **(Figure 2A).** Diabetes type was not associated with overall treatment burden after adjustment (*p*=.46), and the gender difference did not vary by diabetes type (*p*=.25).

**Figure 1.**
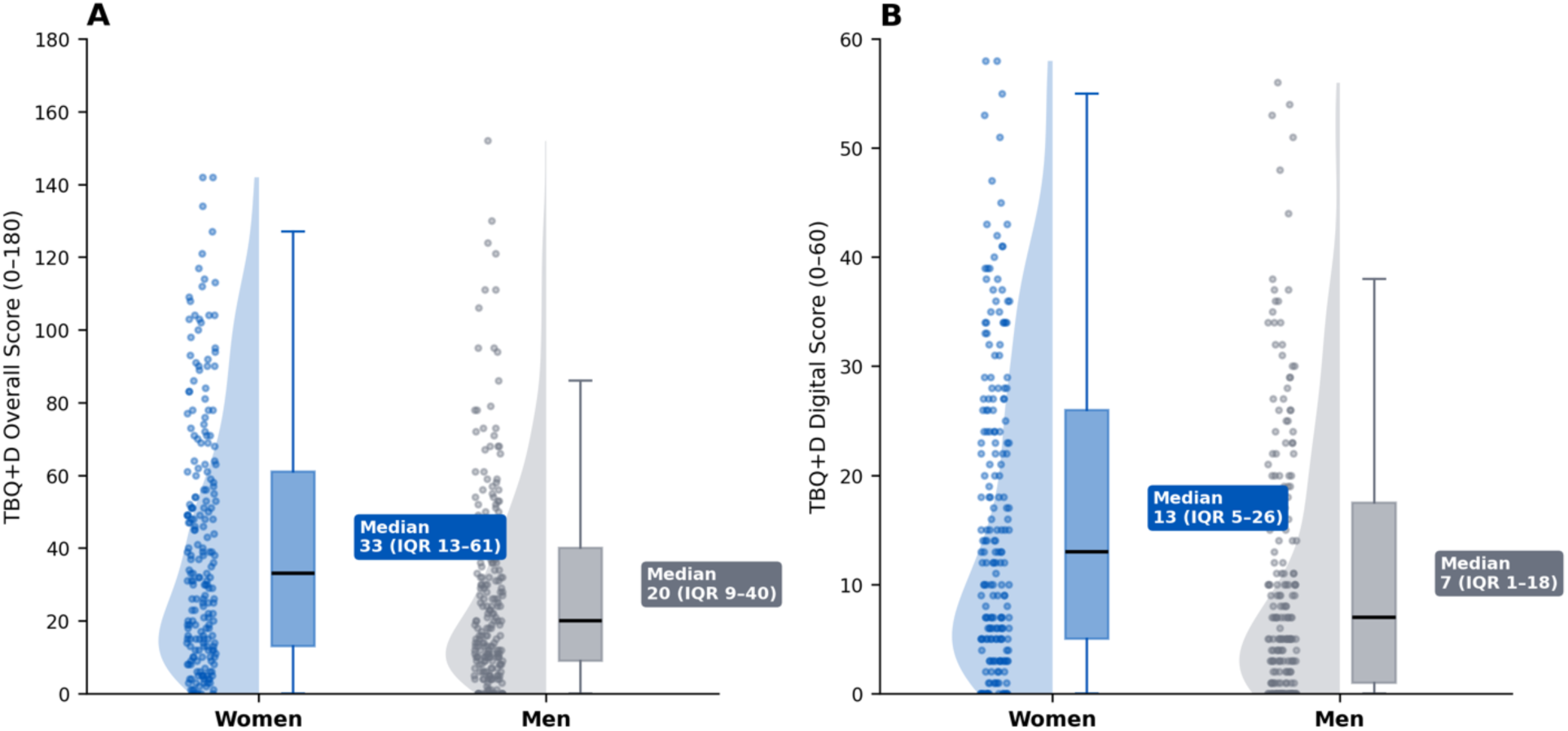
Distribution of overall and digital treatment burden by gender. (A) Overall treatment burden. (B) Digital treatment burden.

**Figure 2.**
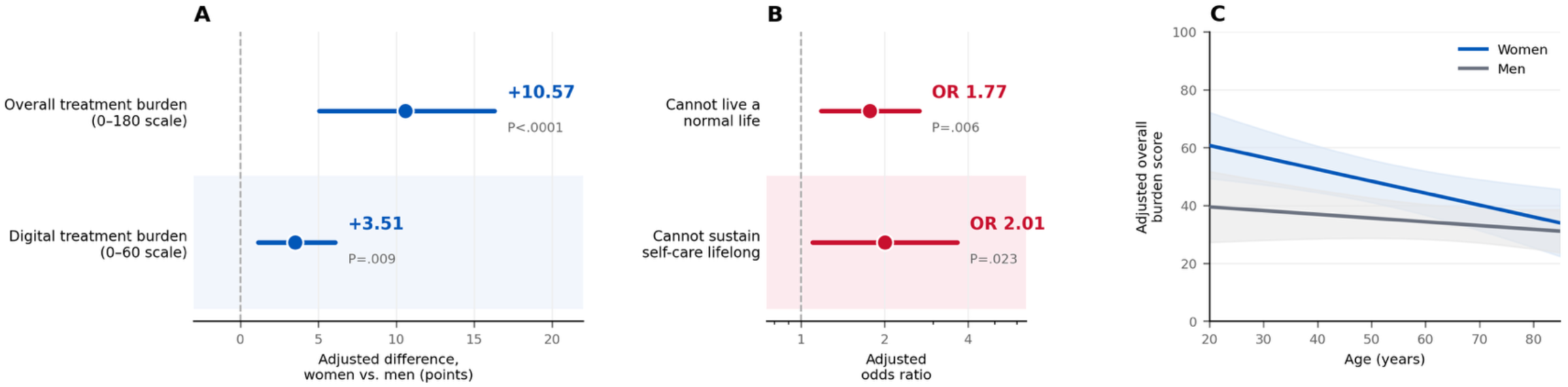
Adjusted gender differences. (A) Overall and digital treatment burden. (B) Inability to live a normal life and difficulty sustaining self-care effort. (C) Overall treatment burden by age and gender. All models were adjusted for age, diabetes type, insulin intensity, HbA1c, and cohort.

Women also had higher adjusted odds of reporting that they were unable to live a normal life (OR 1.77, 95% CI 1.18–2.67; *p*=.006; n=456) and of reporting difficulty sustaining their current level of self-care effort (OR 2.01, 95% CI 1.10–3.66; *p*=.023; n=454) **(Figure 2B).** Overall, 93 (41%) women and 65 (27%) men reported being unable to live a normal life, while 36 (16%) women and 22 (10%) men reported that they could not sustain their current level of self-care effort indefinitely.

Among women, each additional year of age was associated with a 0.36-point lower overall treatment burden score (95% CI −0.64 to −0.09; *p*=.015; n=212), compared with a nonsignificant 0.05-point lower score among men (95% CI −0.25 to 0.16; *p*=.552; n=224). The formal age-by-gender interaction in the pooled Gaussian generalized linear model with log link was not statistically significant (interaction coefficient −0.0049, 95% CI −0.0140 to 0.0042; P=.293; n=436). **(Figure 2C)**.

### Cohort-Stratified Sensitivity Analysis

In cohort-stratified analyses, adjusted gender differences were directionally consistent across the Validation and Encounters cohorts. For overall treatment burden, the adjusted difference between women and men was 13.1 points in the Validation cohort (95% CI 5.9– 20.3) and 7.2 points in the Encounters cohort (95% CI −1.9 to 16.3). For digital treatment burden, the corresponding differences were 3.8 points (95% CI 0.5–7.1) and 3.6 points (95% CI −0.2 to 7.3). The direction of the associations was also consistent for inability to live a normal life and difficulty sustaining the current level of self-care effort (**Supplementary Table 1**).

### Item-Level Drivers of the Gender Gap

The difference in digital treatment burden between women and men was concentrated in a small number of items **(Figure 3)**. Annoyance (21%), precautions (19%), and effort related to using digital medicine tools (18%) were the three largest contributors and together accounted for 59% of the digital burden difference. Need for digital medicine tools also exceeded the equal-share benchmark.

**Figure 3.**
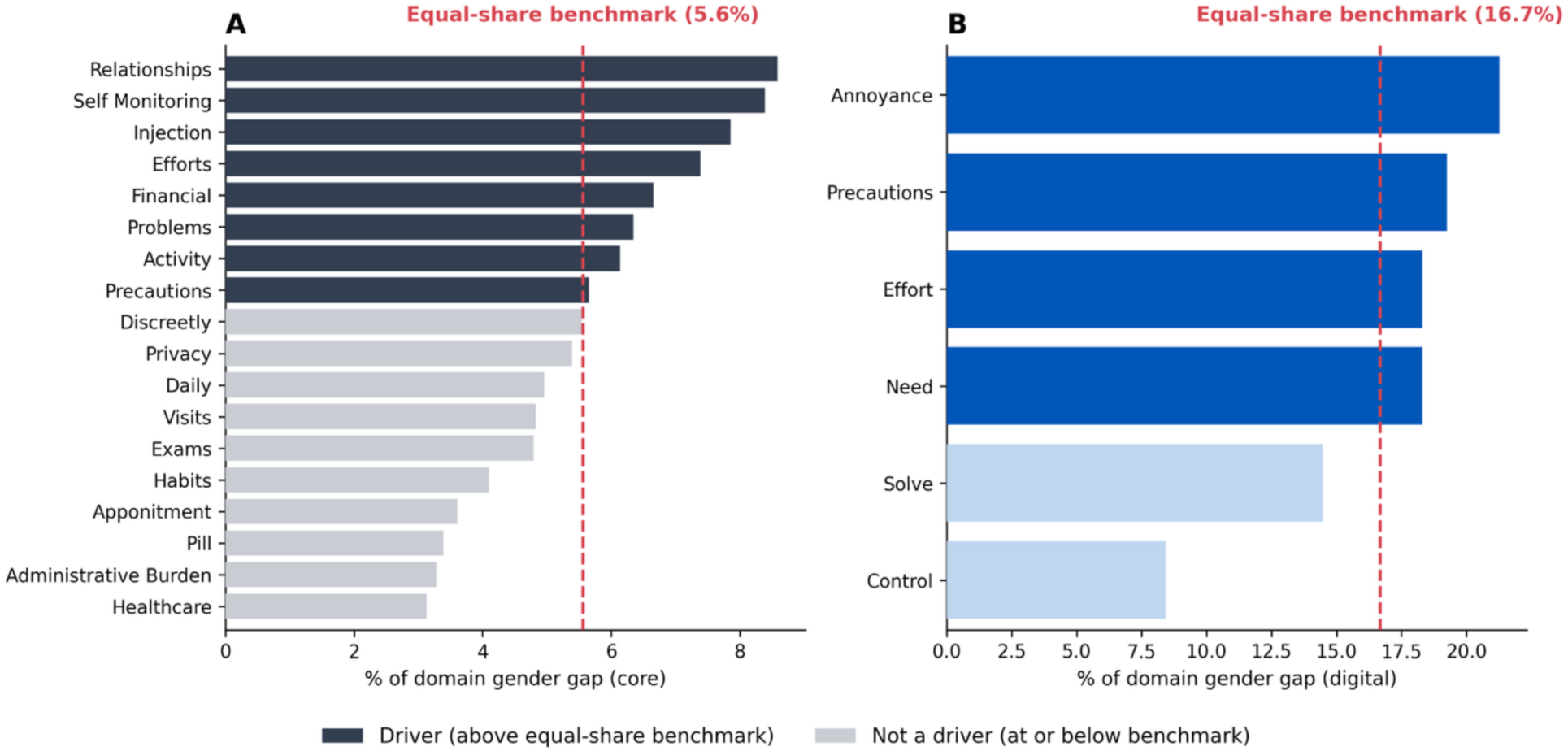
Item-level contributions to the gender difference in treatment burden. (A) Overall treatment burden. (B) Digital treatment burden. Bars show each item’s share of the unadjusted gender difference within its domain. The dashed line marks the equal-share benchmark.

The difference in overall treatment burden was more broadly distributed across items. Relationships (8.6%), self-monitoring (8.4%), and injections (7.9%) were the three largest contributors and together accounted for 25% of the overall treatment burden difference. In adjusted item-level analyses, the overall pattern was similar, although the relative ranking of some items changed. Relationships remained the largest adjusted difference, followed by efforts, problems, self-monitoring, financial burden, and injections. In the digital domain, annoyance remained the largest adjusted item-level difference, followed by need, effort, and precautions **(Supplementary Table 2).**

### Threshold Analysis

Applying the absolute PASS cutoff of 59 points directly to the 18-item overall treatment burden score among the 441 participants with complete scores (213 women and 228 men), 57 women (26.8%) compared with 27 men (11.8%) exceeded the threshold (*p*<.001). Applying the rescaled PASS-based threshold of 70 points, 19% of women compared with 7.5% of men exceeded the threshold (*p*<.001). A sample-specific threshold estimated using the sustainability item as the anchor yielded a cutoff of 44 points; we found that 38% of women and 21% of men exceeded this threshold (*p*<.001).

## CONCLUSIONS

Compared to men, women living with diabetes reported greater overall and digital treatment burden that were more often considered unsustainable and a hindrance to living a normal life while living with diabetes. Although a minimal clinically important difference has not been established for the TBQ+D, an anchor-based analysis offers additional context for the magnitude of the observed difference: nearly two in ten women had treatment burden considered unsustainable compared to 1 in 10 men. These findings, contributed by a broad range of sources of treatment burden, could not be accounted for by differences in treatment intensity and glycemic control.

These findings extend prior evidence of greater treatment burden among women across chronic conditions (7–9) and, to our knowledge, provide the first quantitative evidence of a gender difference in the burden of treatment associated with digital diabetes care. Digital technologies are often intended to facilitate diabetes management, yet they may also create additional work for some patients. The concentration of digital burden in annoyance, precautions, and effort suggests that this work extends beyond access to or use of the technology itself and includes incorporating devices into daily routines and managing the demands they create, consistent with prior qualitative research on digital treatment burden.(2) In contrast, the more broadly distributed pattern in overall treatment burden suggests that no single treatment task accounts for the observed gender difference. The contribution of relationships and self-monitoring is also consistent with qualitative literature describing how caregiving, competing responsibilities, and the social context of illness may shape women’s experience of self-management.(10; 11; 13)

We also observed lower treatment burden with increasing age among women, whereas no significant association with age was observed among men. However, the age-by-gender interaction was not statistically significant, and this finding should therefore be interpreted cautiously. Prior studies have similarly reported greater treatment burden among younger and middle-aged adults than older adults,(24) while research outside diabetes suggests that competing work, caregiving, and family responsibilities may be particularly relevant during these stages of life.(25; 26) Whether these factors explain the observed age pattern will require confirmation in larger studies with direct measures of caregiving, employment, and other demands on patients’ capacity.

These findings have practical implications for diabetes care. Minimally disruptive care calls for clinicians and patients to form plans of care that respond to the patient’s situation by advancing their goals for care and life while minimizing the burden of treatment.(4; 27) Our findings suggest that the burden experienced, predicted to reflect the balance of workload (from having to access and use healthcare and to enact self-care tasks in addition to responding to demands from family, occupation, and community) and capacity (reflecting physical, mental, and financial health, self-efficacy, and social and economic resources) cannot be ascertained by simply analyzing the intensity of treatment (as a measure of the treatment workload) and glycemic control (as a measure of the burden of illness).

Prior work has shown that much of the burden results from systems of healthcare that transfer work to patients and caregivers and are not designed to make accessing and using services easy; these systems are more likely to burden people who are already shouldering work and caregiving responsibilities for others, which current structures preferentially allocate to women.(26; 28) Our findings may also indicate that men experience less burden of care than women in part because women provide capacity to shoulder men’s burden of treatment, as found in a qualitative study of people living with type 2 diabetes in Chile.(29)

Although all patients should receive minimally disruptive care, the TBQ+D can identify patients experiencing significantly high burden of treatment and target them for creative efforts to reduce the treatment workload and to improve patient capacity.(28) These efforts may involve deploying digital tools but our findings demonstrate that some patients may need more support than others for these applications to effectively alleviate, rather than exacerbate, treatment workload.

This study has several limitations. Its cross-sectional design does not establish the direction or mechanisms underlying the observed associations. We were unable to account for diabetes duration or non-diabetes comorbidities, mental health conditions, caregiving responsibilities, and caregiver support, factors which may influence treatment burden. Although unlikely, we did not study potential sex-related contributors to the observed gendered differences. Gender comparisons were limited to women and men because only two participants identified as another gender. Because participants were recruited from a single academic practice, generalizability to other care settings may be limited. Strengths include the use of a validated measure designed specifically to capture both overall and digital treatment burden, replication of the direction and approximate magnitude of the findings across two independently recruited cohorts, and restriction of digital burden analyses to participants who used digital medicine tools.

Compared with men, women living with type 1 or type 2 diabetes experienced greater overall and digital treatment burden and were more likely to be overwhelmed by diabetes care. Assessing treatment burden alongside traditional measures of diabetes control may help identify patients for whom otherwise effective care is becoming difficult to manage and ultimately unsustainable. Attention to treatment workload, including the work created by digital medicine tools, may provide opportunities to better fit diabetes care within patients’ lives.

## Data Availability

The data that support the findings of this study are not publicly available because they contain information that could compromise participant privacy and are subject to institutional and ethical restrictions.

## ACKNOWLEDGMENTS

Personal Thanks

We thank the staff of the Division of Endocrinology, Diabetes, Metabolism, and Nutrition, Department of Medicine, Mayo Clinic, Rochester, Minnesota, for their assistance with participant recruitment and study logistics.

## Funding and Assistance

This study received no external funding.

## Author Contributions and Guarantor Statement

M.A.Z. contributed to conceptualization, methodology, investigation, formal analysis, interpretation of the results, and writing of the original draft. F.D.R. contributed to formal analysis and interpretation of the results. O.M.R. and B.N.S. contributed to data collection.

M.E.B. contributed to methodology, formal analysis, interpretation of the results, and supervision of the statistical analyses. J.P.B. contributed to investigation and interpretation of the results. V.M.M. contributed to conceptualization, methodology, investigation, supervision, and interpretation of the results. All authors contributed to reviewing and editing the manuscript and approved the final version. V.M.M. is the guarantor of this work and, as such, takes responsibility for the integrity of the data and the accuracy of the data analysis.

---

**Supplementary Table 1.**
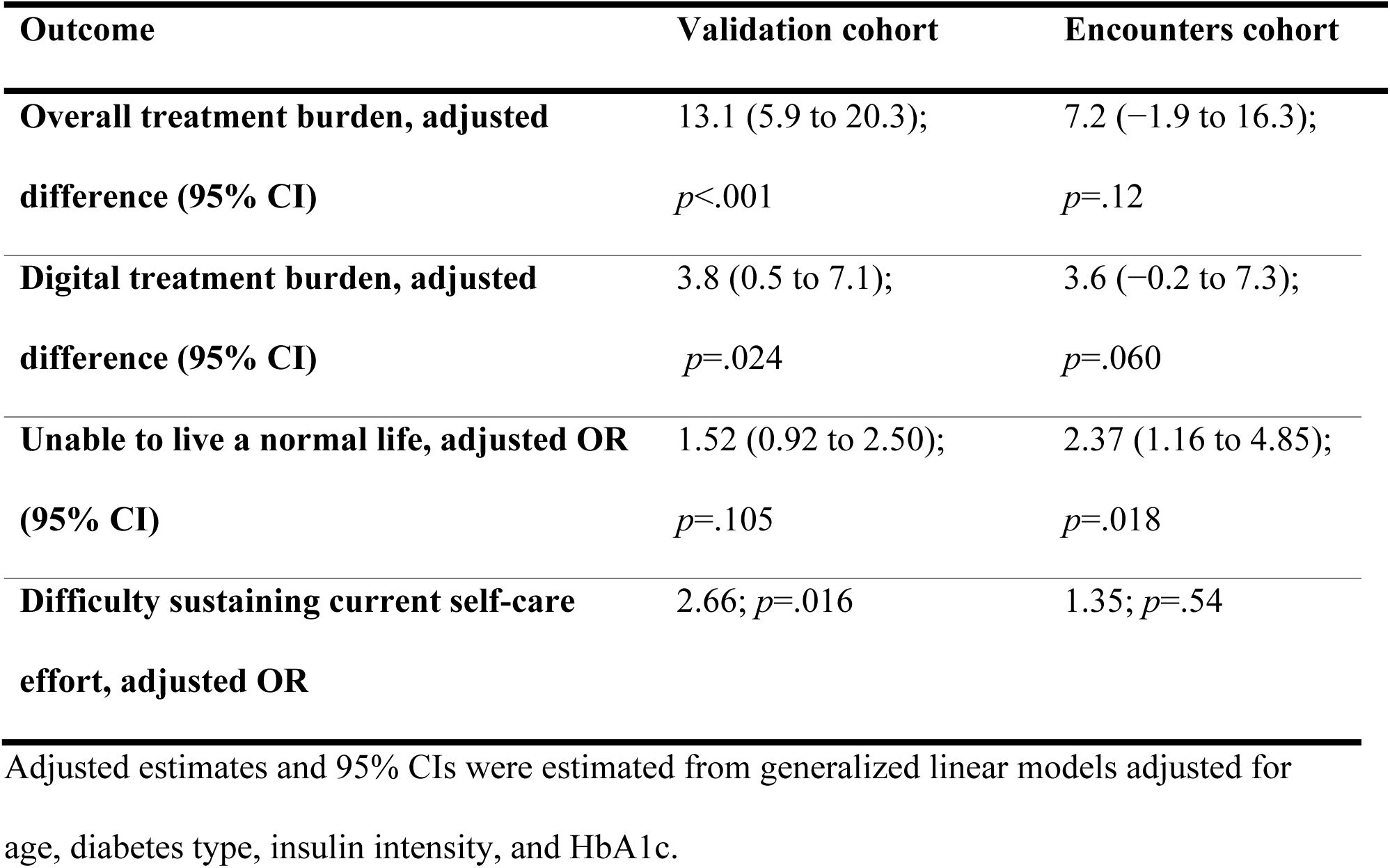
Cohort-stratified adjusted gender differences in treatment burden and care sustainability.

**Supplementary Table 2.**
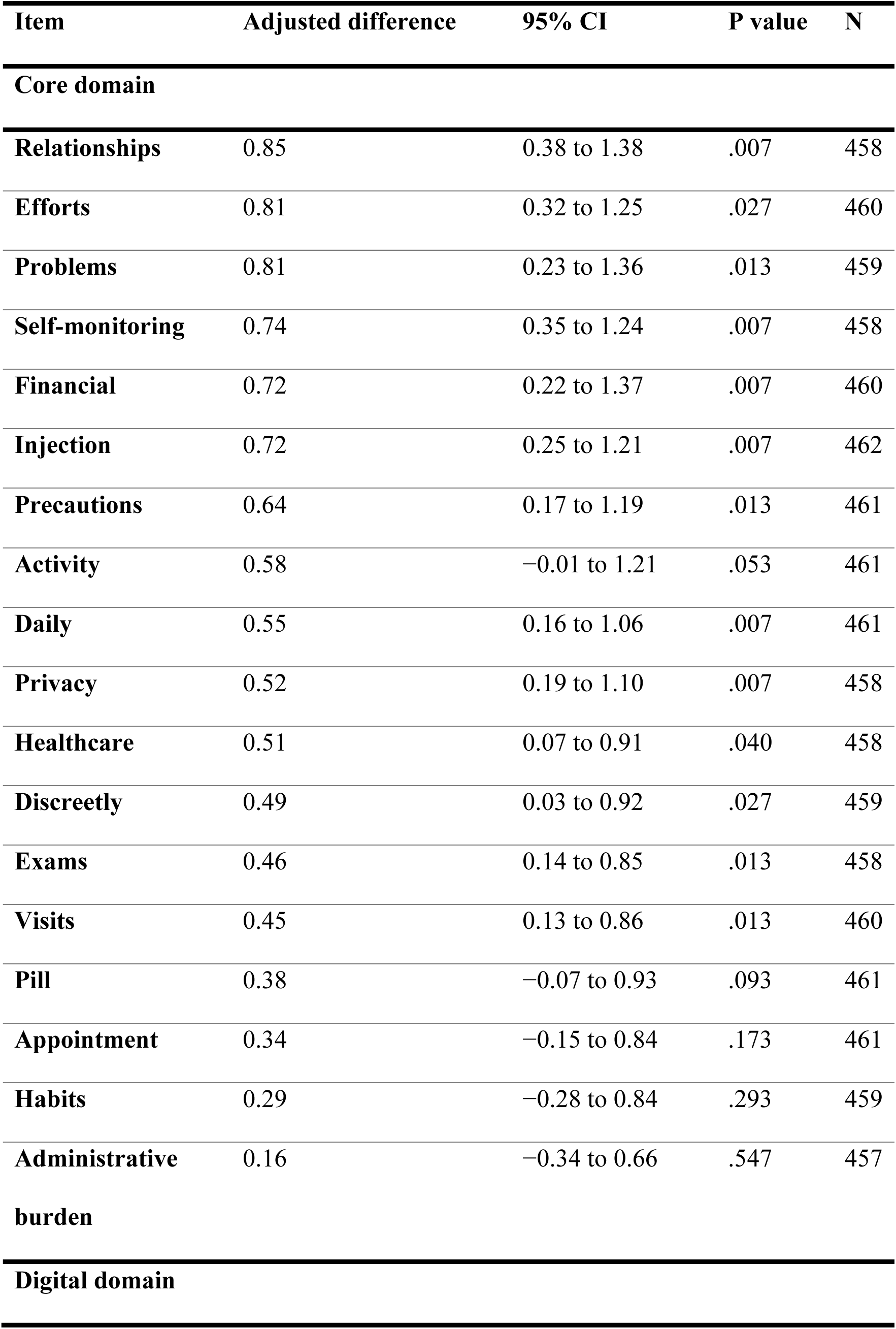
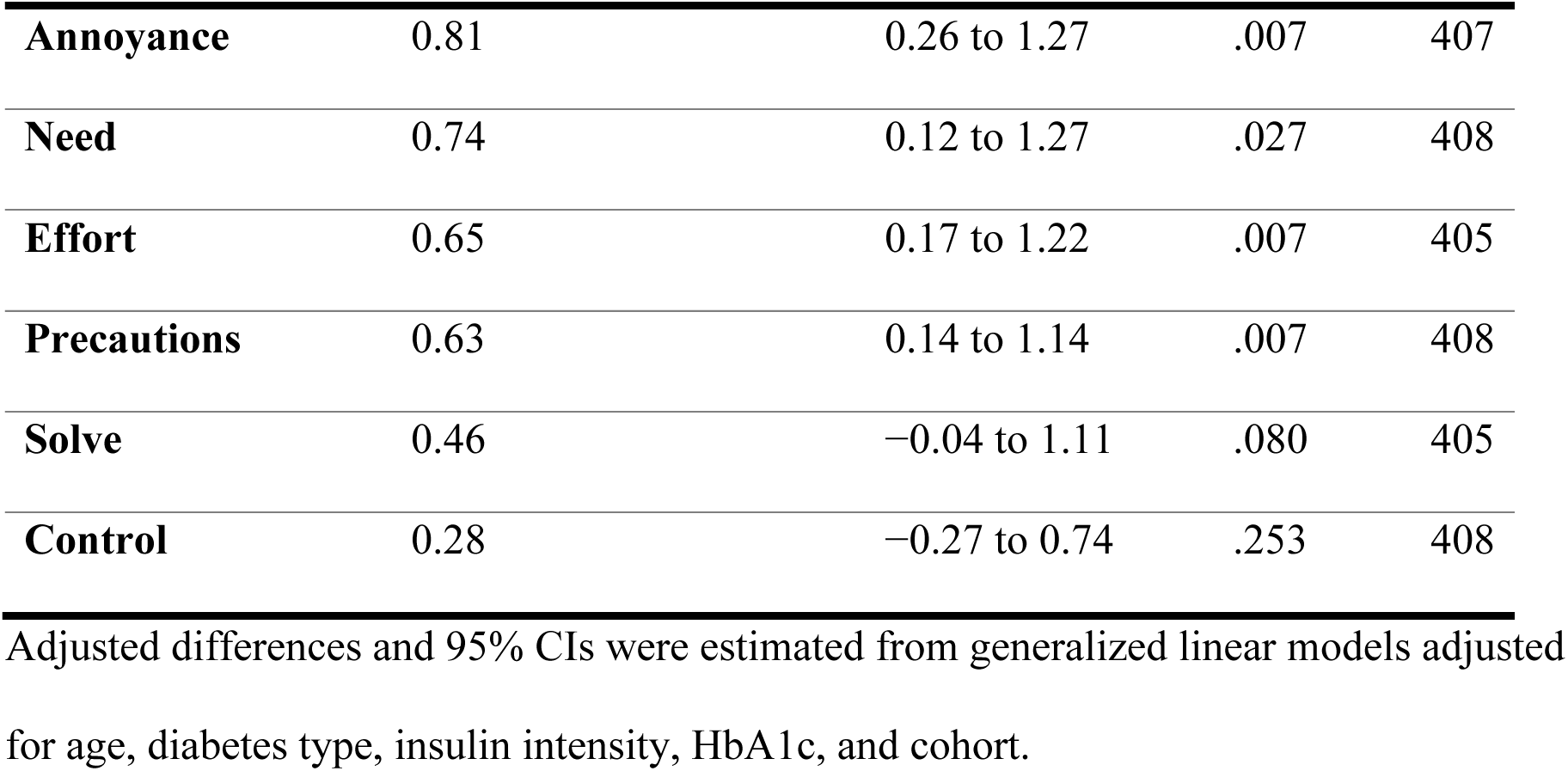
Adjusted item-level gender differences in TBQ+D scores.

